# Comparing the Cost-Effectiveness of Alternative Policies for Recommending and Providing HIV Pre-exposure Prophylaxis to Men Who Have Sex With Men in the EU

**DOI:** 10.1101/2025.01.28.25321249

**Authors:** Boxuan Wang, João Brázia, Andreia Sofia Teixeira, Eugenio Valdano

**Affiliations:** Sorbonne Université, INSERM, Institut Pierre Louis d’Epidémiologie et de Santé Publique, F75012, Paris, France; LASIGE, Departamento de Informática, Faculdade de Ciências, Universidade de Lisboa, Lisbon, Portugal; Network Science Institute, Northeastern University London, London, E1W 1LP, United Kingdom

## Abstract

**Background:** In the European Union (EU), HIV disproportionately affects men who have sex with men (MSM), with prevalence rates ranging from 2.4% to 29%. Despite the high efficacy of pre-exposure prophylaxis (PrEP) in preventing HIV, its accessibility and uptake remain uneven across **countries**. This study examines the effectiveness and cost-effectiveness of different PrEP policies across 20 EU countries.

**Methods:** We employed a stochastic agent-based model of HIV transmission among MSM. The model incorporated data on sexual behavior, PrEP adherence and healthcare costs to evaluate the impact of five distinct PrEP eligibility policies. Outcomes included HIV infections averted, HIV-related deaths prevented, PrEP coverage, and incremental cost-effectiveness ratio.

**Findings:** Policies by the US, CDC, and Belgian authorities were the most effective in reducing HIV infections and deaths, driven by higher PrEP coverage. However, the WHO policy emerged as the most cost-effective across the 20 countries, despite its current use being limited to Denmark. The European AIDS Clinical Society guidelines also showed potential, although not currently implemented. In countries without PrEP reimbursement, reducing drug costs would significantly expand the range of cost-effective policy options.

**Interpretation:** Optimizing PrEP policies is crucial for reducing HIV incidence among MSM in the EU. Broad eligibility criteria maximize effectiveness, while WHO guidelines offer a cost-effective alternative in constrained economic contexts. Our findings highlight the need for policy adjustments to enhance PrEP accessibility, inform national health strategies, and achieve sustainable HIV prevention across diverse settings.

**Funding:** Campus France PHC Pessoa, French Government; Portugal-France Bilateral Co-operation 2022 Programa Pessoa ref. 2022.15068.CBM; FCT -- Fundação para a Ciência e Tecnologia -- through the LASIGE Research Unit, ref. UIDB/00408/2020 and ref. UIDP/00408/2020; Horizon Europe grant SIESTA (101131957).

## Introduction

Countries in the European Union (EU) have achieved substantial progress in the fight against the HIV/AIDS epidemic in the last decades. However, men who have sex with men (MSM) are still disproportionally affected, and intersecting vulnerabilities exacerbate this: it is the case of migrant MSM who exhibit high rate of acquisition following migration to Europe(1). Improving primary prevention among MSM has been identified as a crucial step to avert infections in this population(2). This involves pre-exposure prophylaxis (PrEP), the use of antiretroviral medication to prevent HIV acquisition even during unprotected sex. PrEP has been shown to be safe and effective in preventing HIV acquisition(3–7), but its availability and uptake is highly heterogeneous across European countries. Barriers to the scale-up of PrEP include concerns over its possible negative effects on the circulation of bacterial sexually transmitted infections(8) and their rates of antibiotic resistance(9). At the same time, evidence in support of these concerns is mixed(10) and these barriers might themselves reinforce stigma(11) and hamper comprehensive prevention strategies(12). As a result of these opposing arguments, countries in the EU have made starkly different choices on how much to invest on, make available, and encourage PrEP among MSM. Countries like France fully reimburse PrEP and recommend it to all those who identify as gay men or report having sex with men. Others, like Belgium, reimburse it but prescribe it only to those with an estimated high risk of being exposed to HIV, and only through specialized visits in national healthcare centers or hospitals. In others, like Hungary and Italy, PrEP is not reimbursed but it is available upon prescription. Finally, in others like Romania, PrEP is not available(13).

A crucial determinant of the different PrEP policies is their definition of eligibility. Eligibility criteria vary across guidelines and generally rely on estimating the risk of HIV exposure based on socio-demographic factors and sex-related behaviors. These behaviors include reported condom use, being in a serodiscordant partnership, the use of postexposure prophylaxis, a history of sexually transmitted infections, or the use of psychoactive substances during sex. This results in substantially different pools of eligible individuals across different guidelines, with substantially different risks of acquiring HIV(14,15).

This heterogeneity of approaches across EU countries highlights the need for an assessment of PrEP effectiveness and cost-effectiveness across healthcare systems and policy guidelines. Currently, most studies assessing the cost-effectiveness of PrEP policies in high-income countries focus on the United States(16). Assessments of the cost and cost-effectiveness of PrEP in the EU are scarce, and focus on single countries(17–20). Results are varied across countries and settings, indicating that PrEP may be cost-effective only under certain conditions, like reduced drug price or event-driven use(21). Some studies recommended providing PrEP only to high-risk MSM, while others found that risk-based eligibility may reinforce stigma and decrease the overall effectiveness of the prevention campaign(22,23). Crucially, two elements are still missing: to systematically evaluate the cost-effectiveness of the PrEP policies that each country is adopting, and to determine the potential benefits of switching to an alternative policy. To address these knowledge gaps, in this study, we focused on ten countries in the EU which currently provide free PrEP to MSM. We estimated the impact and cost-effectiveness of their current PrEP policies on HIV infections and related deaths over the next decades. We also tested the hypothetical implementation of alternative policies among those recommended by a set of EU and non-EU authorities. Additionally, we studied ten other EU countries which currently have no free PrEP policy, and tested the impact and cost-effectiveness of the different policies. We employed a previously developed stochastic, agent-based model(24) of HIV infection among MSM, and used Incremental Cost-Effectiveness Ratio (ICER) to determine cost-effectiveness(25).

## Methods

### Countries and PrEP guidelines under study

We considered the following countries of the EU which currently provide PrEP free of charge: Belgium, Denmark, Finland, France, Germany, Ireland, Netherlands, Portugal, Slovenia, Spain. We also considered these countries which do not currently reimburse PrEP: Bulgaria, Greece, Hungary, Italy, Latvia, Lithuania, Poland, Cyprus, Malta, Romania. We tested five different PrEP eligibility guidelines, labeled according to the organization/institute recommending it or the country adopting it. The most restrictive is POR, by the Portuguese Ministry of Health(14). It recommends prescribing PrEP to those reporting unprotected anal sex and whose partner has not tested for HIV in the past six months. In addition to Portugal, it is currently in use in Finland, Ireland, Netherlands, and Slovenia. It should be noted that, henceforth, *in use* just means that equivalent guidelines are in place, not that another country’s guidelines are explicitly adopted. The second most restrictive is the World Health Organization’s Implementation Tool for Pre-exposure Prophylaxis of HIV Infection (WHO)(26), which recommends PrEP to those declaring unprotected anal sex and have more than one sex partner. It is currently in use in Denmark. The third one is the European AIDS Clinical Society’s Guidelines Version 9 (EACS)(27) which recommends PrEP to those reporting unprotected anal sex in non-main partnerships. The least restrictive are the United States Centers for Disease Control and Preventions (CDC) Public Health Service(28), which recommends PrEP to all those identifying as MSM and is currently in use in France and Germany, and that of the Belgian Federal Office of Health (BEL). BEL recommends PrEP to all those reporting unprotected anal sex(29) and is currently in use in Belgium and Spain. See Figure 1 for a map of the countries under study, the situation of their HIV epidemic among MSM, and the PrEP policy now in effect.

**Figure 1:**
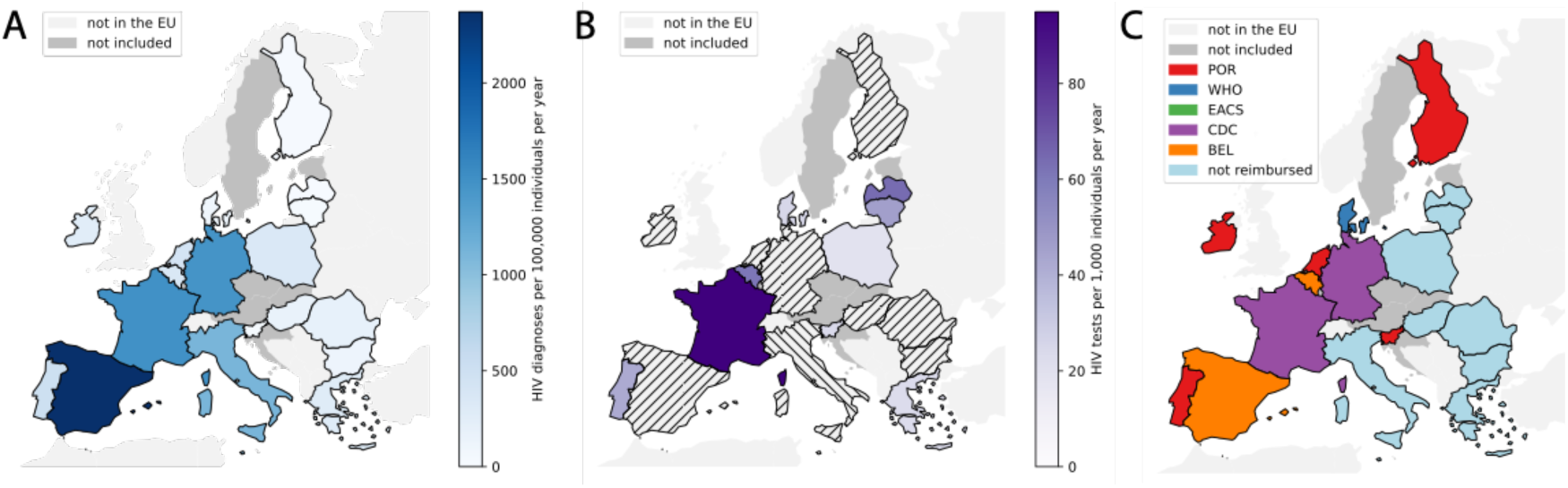
Countries considered in the study. A) Rate of HIV diagnoses among men, with acquisition occurring through sex with men, per year, per 100,000 individuals. Data from 2019 (ECDC). B) Number of HIV tests performed, except anonymous tests, per year, per 1,000 individuals. Data from 2022 (ECDC). Hashing indicates that data were not available. C) Currently adopted PrEP policies in each country.

### Transmission model and PrEP dynamics

We built a network-based, stochastic agent-based model of HIV transmission within the EpiModel framework (version 1.2.5), developed in Ref.(24). EpiModel has already been used to model the impact of PrEP on the HIV epidemics(30,31). In our study, we adopted a weekly time scale and a synthetic population of 10,000 men. The model features a multilayer network encoding primary, casual and one-time partnerships, implemented using exponential random graph models(32). The model accommodated individual characteristics such as ethnicity, circumcision status, sexual preference (receptive/insertive), and propensity for unprotected anal sex. The model was parametrized using the EMIS cross-sectional study(33), the Lisbon Cohort study (34) and Ref(35) to inform partnership network dynamics (see Appendix pp 2-3 for further details). Synthetic population dynamics consists in male individuals entering the population at 18 years of age and exiting at death or when they become 45 years old. In each country, the model was initialized at the rate of HIV diagnoses among MSM as recorded by EMIS(33) (Figure 1). After a transient that was discarded to exclude effects due to the initial seeding, the model was then run for 20 years into the future. For each model set-up, 1,000 runs are performed and observables of interest are measured as median and 95% confidence intervals across runs. Additional details on the agent-based model are available in the Appendix pp 3-6. We assumed a 20% probability of an eligible individual initiating PrEP in any given week, based on previous studies from high-income countries(34).

### Health economics analysis

In the countries where PrEP is reimbursed, we adopted a payer/direct cost perspective. To estimate the annual cost of PrEP, we used the official minimum reimbursable price of PrEP in each country as the cost of PrEP(13). This estimation does not include costs associated with regular testing or monitoring, allowing us to concentrate on the direct costs of the PrEP intervention. We used the ECDC progress report(36) for the cost of antiretroviral therapy (ART) in each country. For the cost-effectiveness analysis, we used Quality-Adjusted Life Year (QALY). QALY measures the quality and quantity of life lived and is commonly used in health economics and cost-effectiveness analyses. We used a utility value of 0.7 for HIV infection, regardless of its stage(37). We then computed the Incremental Cost-Effectiveness Ratio (ICER) which is the additional cost per QALY gained. As customary, we set the threshold of cost-effectiveness to twice the gross domestic product (GDP) per capita, based on World Bank data(38). In the countries that reimburse PrEP, a policy was considered cost-effective if its ICER was below that threshold. We also compared different policies according to their ICER, to determine their relative cost-effectiveness. In the countries that do not reimburse PrEP, we used the cost-effectiveness threshold to determine the willingness-to-pay threshold. The willingness-to-pay threshold is the highest cost of PrEP at which the policy would still be cost-effective. We set a yearly discount rate of 0 %. In sensitive analysis, yearly discount rate of 2 % has been considered. This report has been prepared following the Consolidated Health Economic Evaluation Reporting Standards (CHEERS) guidelines(39). The completed CHEERS checklist is provided in the Appendix pp 11-12.

## Results

### Policy effectiveness on infections, deaths and PrEP coverage

We first focused on countries that currently reimburse PrEP. Expectedly, stringent policies led to a long-term PrEP coverage that was substantially lower than that of less stringent policies. Coverage values are reported in Tables 1 and 2. We estimated that after 20 years, POR – the most stringent policy – achieved a PrEP coverage of around 48% across countries almost half of what we estimated that the CDC policy would achieved -- almost 90%. Moreover, we observed that PrEP coverage stabilized across countries and policies within the first 10 years of implementation (compare PrEP coverage after 10 – Table 1 - and 20 years - Table 2). As expected, we observed that all policies led to a substantial decrease in HIV infections and HIV-related deaths in the next 20 years, as compared to the counterfactual scenario of no PrEP (see Appendix pp 7). Their effectiveness, however, was heterogeneous. Tables 1 and 2 report the rates of averted infections and deaths across countries and guidelines, and Figure 2 ranks them by their effectiveness. We can see that the CDC policy was the most effective in averting infections over the first 10 years in every country, and that the EACS policy ranked second in 9 out of 10 countries. These policies averted, respectively, 76-to-142 and 68-to-134 infections per 1,000 individuals. The BEL and CDC policies were instead the most effective in averting deaths: 15-24 and 13-27 per 1,000 individuals, respectively. BEL also had substantial impact on averted infections, ranging from 63 to 129 per 1,000 individuals. Notably, the impact of EACS on averting deaths was negligible. WHO and POR, the two most restrictive policies, had a comparable impact on averting infections, albeit strongly country-dependent: 52-to-100 and 33-to-71, respectively. Their impact on averting deaths was substantially stronger than EACS: 7-to-17 and 8-to-20, respectively.

**Figure 2 :**
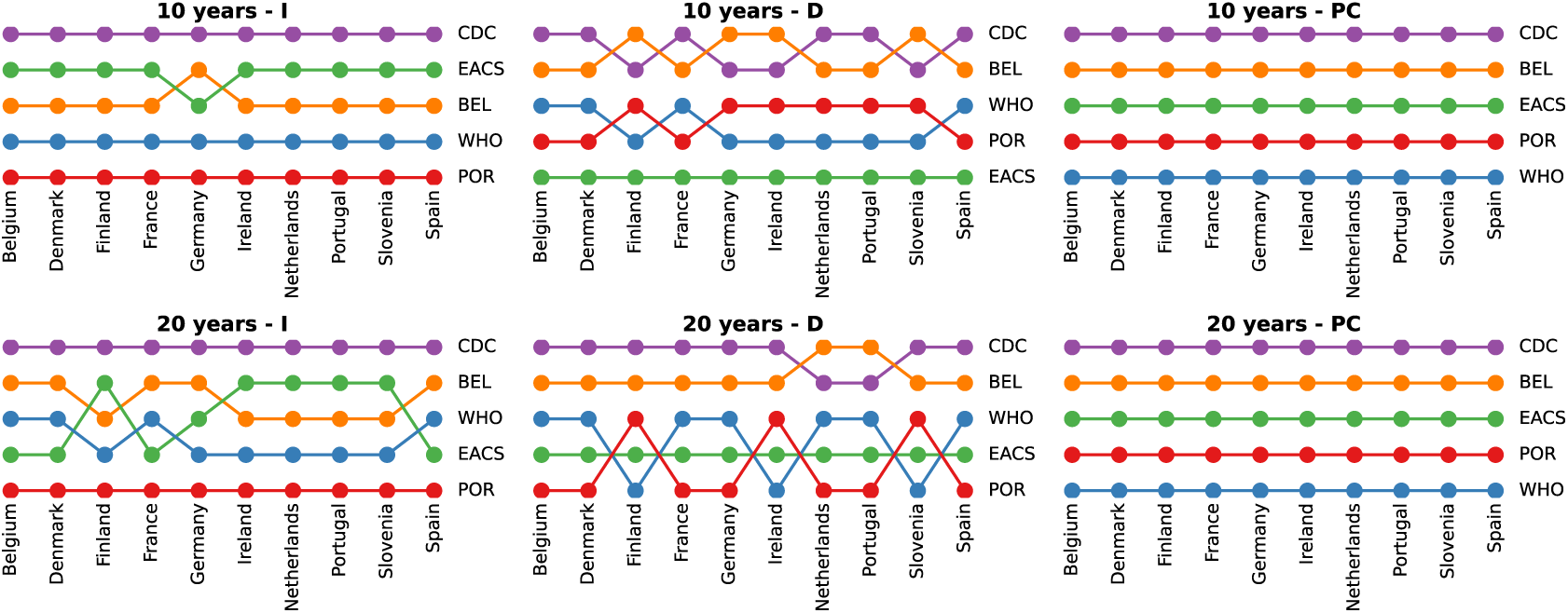
Ranking of PrEP policies in each country according to their effectiveness in averting HIV infections (I), in averting HIV-related deaths (D) and in achieving high PrEP coverage (PC). Rankings evaluated over 10 and 20 years. See Table 1 for the estimated effectiveness values.

**Table 1:** Number of averted HIV infections (I) and deaths (D) per 1,000 individuals, and PrEP coverage (PC, percent %), in the countries that currently reimburse PrEP, under the different PrEP policies, at the 10-year time horizon. The reported values are median and 95% confidence interval from the stochastic iterations of the model.

| Time period: 10 years |  |  |  |  |  |  |  |  |  |  |  |  |  |  |  |
| --- | --- | --- | --- | --- | --- | --- | --- | --- | --- | --- | --- | --- | --- | --- | --- |
|  | EACS |  |  | CDC |  |  | WHO |  |  | POR |  |  | BEL |  |  |
|  | I | D | PC | I | D | PC | I | D | PC | I | D | PC | I | D | PC |
| **Belgium** | 114(93-120) | 3 (-1-6) | 68 (67-69) | 127(123-135) | 27 (22-32) | 88 (88-89) | 93(90-95) | 16 (6-16) | 39 (37-40) | 61(55-65) | 14 (13-18) | 48 (46-49) | 98(97-118) | 18 (15-21) | 74 (72-74) |
| **Denmark** | 114(93-120) | 3 (-1-6) | 68 (67-69) | 127(123-135) | 27 (22-32) | 88 (88-89) | 93(90-95) | 16 (6-16) | 39 (37-40) | 61(55-65) | 14 (13-18) | 48 (46-49) | 98(97-118) | 18 (15-21) | 74 (72-74) |
| **Finland** | 68(45-93) | 0 (-4-2) | 64 (63-66) | 76(70-90) | 13 (13-18) | 85 (84-86) | 52(49-74) | 7 (7-8) | 35 (33-37) | 33(30-51) | 9 (7-11) | 43 (42-45) | 63(54-92) | 15 (14-16) | 69 (68-70) |
| **France** | 114(93-120) | 3 (-1-6) | 68 (67-69) | 127(123-135) | 27 (22-32) | 88 (88-89) | 93(90-95) | 16 (6-16) | 39 (37-40) | 61(55-65) | 14 (13-18) | 48 (46-49) | 98(97-118) | 18 (15-21) | 74 (72-74) |
| **Germany** | 104(87-105) | 6 (5-7) | 65 (64-66) | 127(98-127) | 24 (24-25) | 86 (84-86) | 96(79-105) | 17 (16-18) | 37 (36-40) | 71(68-73) | 20 (18-22) | 45 (43-46) | 105(100-110) | 24 (15-29) | 70 (68-71) |
| **Ireland** | 68(45-93) | 0 (-4-2) | 64 (63-66) | 76(70-90) | 13 (13-18) | 85 (84-86) | 52(49-74) | 7 (7-8) | 35 (33-37) | 33(30-51) | 9 (7-11) | 43 (42-45) | 63(54-92) | 15 (14-16) | 69 (68-70) |
| **Netherlands** | 134(100-136) | 2 (-3-7) | 66 (65-67) | 142(128-152) | 27 (23-28) | 86 (86-87) | 100(88-107) | 8 (6-9) | 37 (36-39) | 55(43-66) | 8 (7-9) | 46 (44-47) | 129(89-136) | 24 (10-21) | 71 (70-72) |
| **Portugal** | 134(100-136) | 2 (-3-7) | 66 (65-67) | 142(128-152) | 27 (23-28) | 86 (86-87) | 100(88-107) | 8 (6-9) | 37 (36-39) | 55(43-66) | 8 (7-9) | 46 (44-47) | 129(89-136) | 24 (10-21) | 71 (70-72) |
| **Slovenia** | 68(45-93) | 0 (-4-2) | 64 (63-66) | 76(70-90) | 13 (13-18) | 85 (84-86) | 52(49-74) | 7 (7-8) | 35 (33-37) | 33(30-51) | 9 (7-11) | 43 (42-45) | 63(54-92) | 15 (14-16) | 69 (68-70) |
| **Spain** | 114(93-120) | 3 (-1-6) | 68 (67-69) | 127(123-135) | 27 (22-32) | 88 (88-89) | 93(90-95) | 16 (6-16) | 39 (37-40) | 61(55-65) | 14 (13-18) | 48 (46-49) | 98(97-118) | 18 (15-21) | 74 (72-74) |

**Table 2:** Number of averted HIV infections (I) and deaths (D) per 1,000 individuals, and PrEP coverage (PC, percent %), in the countries that currently reimburse PrEP, under the different PrEP policies, at the 20-year time horizon. The reported values are median and 95% confidence interval from the stochastic iterations of the model.

| Time period: 20 years |  |  |  |  |  |  |  |  |  |  |  |  |  |  |  |
| --- | --- | --- | --- | --- | --- | --- | --- | --- | --- | --- | --- | --- | --- | --- | --- |
|  | EACS |  |  | CDC |  |  | WHO |  |  | POR |  |  | BEL |  |  |
|  | I | D | PC | I | D | PC | I | D | PC | I | D | PC | I | D | PC |
| Belgium | 107<br>(151-161) | 45 (37-61) | 67 (67-68) | 183<br>(175-194) | 65 (57-81) | 90 (91-91) | 148<br>(144-153) | 52 (49-71) | 37 (36-39) | 100 (99-101) | 41 (38-58) | 48 (47-50) | 161<br>(153-172) | 61 (51-70) | 75 (74-75) |
| Denmark | 107<br>(151-161) | 45 (37-61) | 67 (67-68) | 183<br>(175-194) | 65 (57-81) | 90 (91-91) | 148<br>(144-153) | 52 (49-71) | 37 (36-39) | 100 (99-101) | 41 (38-58) | 48 (47-50) | 161<br>(153-172) | 61 (51-70) | 75 (74-75) |
| Finland | 97<br>(82-104) | 30 (20-33) | 65 (64-66) | 112 (89-121) | 45 (32-37) | 89 (89-90) | 84 (81-89) | 29 (21-39) | 35 (34-37) | 65 (63-68) | 32 (26-38) | 44 (43-46) | 91 (91-111) | 39 (31-47) | 72 (71-73) |
| France | 107<br>(151-161) | 45 (37-61) | 67 (67-68) | 183<br>(175-194) | 65 (57-81) | 90 (91-91) | 148<br>(144-153) | 52 (49-71) | 37 (36-39) | 100 (99-101) | 41 (38-58) | 48 (47-50) | 161<br>(153-172) | 61 (51-70) | 75 (74-75) |
| Germany | 139<br>(128-139) | 42 (27-53) | 66 (66-67) | 162<br>(143-159) | 60 (40-64) | 89 (89-90) | 129<br>(114-136) | 50 (45-53) | 37 (35-39) | 94 (94-95) | 40 (36-54) | 46 (44-47) | 147<br>(135-152) | 55 (43-66) | 72 (72-74) |
| Ireland | 97<br>(82-104) | 30 (20-33) | 65 (64-66) | 112 (89-121) | 45 (32-37) | 89 (89-90) | 84 (81-89) | 29 (21-39) | 35 (34-37) | 65 (63-68) | 32 (26-38) | 44 (43-46) | 91 (91-111) | 39 (31-47) | 72 (71-73) |
| Netherlands | 176<br>(156-171) | 42 (34-48) | 66 (66-67) | 197<br>(174-198) | 56 (49-63) | 90 (90-90) | 138<br>(132-147) | 42 (38-47) | 37 (35-38) | 96 (84-108) | 38 (36-40) | 46 (45-48) | 166<br>(157-179) | 56 (50-62) | 73 (73-74) |
| Portugal | 176<br>(156-171) | 42 (34-48) | 66 (66-67) | 197<br>(174-198) | 56 (49-63) | 90 (90-90) | 138<br>(132-147) | 42 (38-47) | 37 (35-38) | 96 (84-108) | 38 (36-40) | 46 (45-48) | 166<br>(157-179) | 56 (50-62) | 73 (73-74) |
| Slovenia | 97<br>(82-104) | 30 (20-33) | 65 (64-66) | 112 (89-121) | 45 (32-37) | 89 (89-90) | 84 (81-89) | 29 (21-39) | 35 (34-37) | 65 (63-68) | 32 (26-38) | 44 (43-46) | 91 (91-111) | 39 (31-47) | 72 (71-73) |
| Spain | 107<br>(151-161) | 45 (37-61) | 67 (67-68) | 183<br>(175-194) | 65 (57-81) | 90 (91-91) | 148<br>(144-153) | 52 (49-71) | 37 (36-39) | 100 (99-101) | 41 (38-58) | 48 (47-50) | 161<br>(153-172) | 61 (51-70) | 75 (74-75) |

Considering the larger time horizon of 20 years, CDC and BEL were the most effective policies across the board. Their rates of averted infections and deaths roughly doubled from 10 to 20 years, indicating that they had achieved their maximum effectiveness by 10 years. EACS instead showed a limited increase in averted infections with respect to the 10-year horizon, bringing down its effectiveness ranking in 5 of the countries in which used to rank second. But its effectiveness in averting deaths substantially improved (30-to-45 for 1,000 individuals). WHO kept the same performance as after 10 years, ranked fourth in terms of averted infections and third or fourth in terms of averted deaths. Overall, rates broadly doubled with respect to the 10-year window, again indicating that the policies had achieved its maximum effectiveness after 10 years. POR, too, roughly doubled the rate of averted infections and deaths from the 10-year window. However, it still was the policy leading to the smallest number of averted infections and ranked third in terms of averted deaths.

In countries that currently do not reimburse PrEP, coverage stabilized between 35% (WHO) and 88% (CDC) across all policies by year 10, with minimal increases observed by year 20 (see Appendix pp 9- 10). PrEP implementation led to substantial reductions in HIV infections and HIV-related deaths compared to the counterfactual scenario of no PrEP. At the 20-year window, the rates of averted infections were more heterogeneous across these countries than across those which reimburse PrEP. This is visible for the CDC policy, which ranked best with 56-to-148 averted infections per 1,000 individuals, as well as for the POR policy, which had the smallest impact (44-to-100). The ranking of intermediate policies (WHO, EACS, and BEL) varied by country: in Cyprus and Latvia, for instance, BEL outperformed WHO which in turn outperformed EACS, whereas in Italy EACS outperformed BEL which in turn outperformed WHO (see Appendix pp 11).

### Health economics analysis in countries reimbursing PrEP

The cost-effectiveness threshold, set at twice the GDP per person, was heterogeneous across the countries which currently reimburse PrEP, ranging from 212,118 euros per QALY in Ireland to 57,938 euros per QALY in Portugal (Appendix pp 9 reports all the cost-effectiveness thresholds). The cost of PrEP was also heterogeneous(13) (see Appendix pp 8-9). We evaluated the cost-effectiveness of currently implemented policies in the countries that reimburse PrEP at the time horizons of 10 and 20 years (Tables 3 and 4). Also, we estimated the cost-effectiveness of all possible alternatives, under the assumption that countries could switch from the one they are currently implementing to any other. Figure 3A reports Incremental Cost-Effectiveness Ratio (ICER) at the time horizon of 10 years and shows that, the currently implemented policies were cost-effective only in Denmark (policy WHO), the Netherlands (POR) and Spain (BEL). It is possible to also observe that the WHO policy would be cost-effective if implemented in Belgium, Germany, the Netherlands, Portugal and Spain. The EACS policy would be cost-effective, if implemented, in Germany, Ireland, the Netherlands, Portugal and Spain. The CDC policy would be cost-effective in Ireland, the Netherlands, Portugal and Spain. Finally, the BEL policy would be cost-effective in the Netherlands and Portugal. Notably, all policies would be cost-effectiveness in the Netherlands, while none would be cost-effective in Finland and Slovenia. The currently implemented policy was the most cost-effectiveness only in Denmark (WHO). In all the other countries, which reimbursed PrEP, the most cost-effective policy was either WHO (Belgium, France, Germany) or EACS (Finland, Ireland, the Netherlands, Portugal, Slovenia – tied with CDC – and Spain).

**Figure 3.**
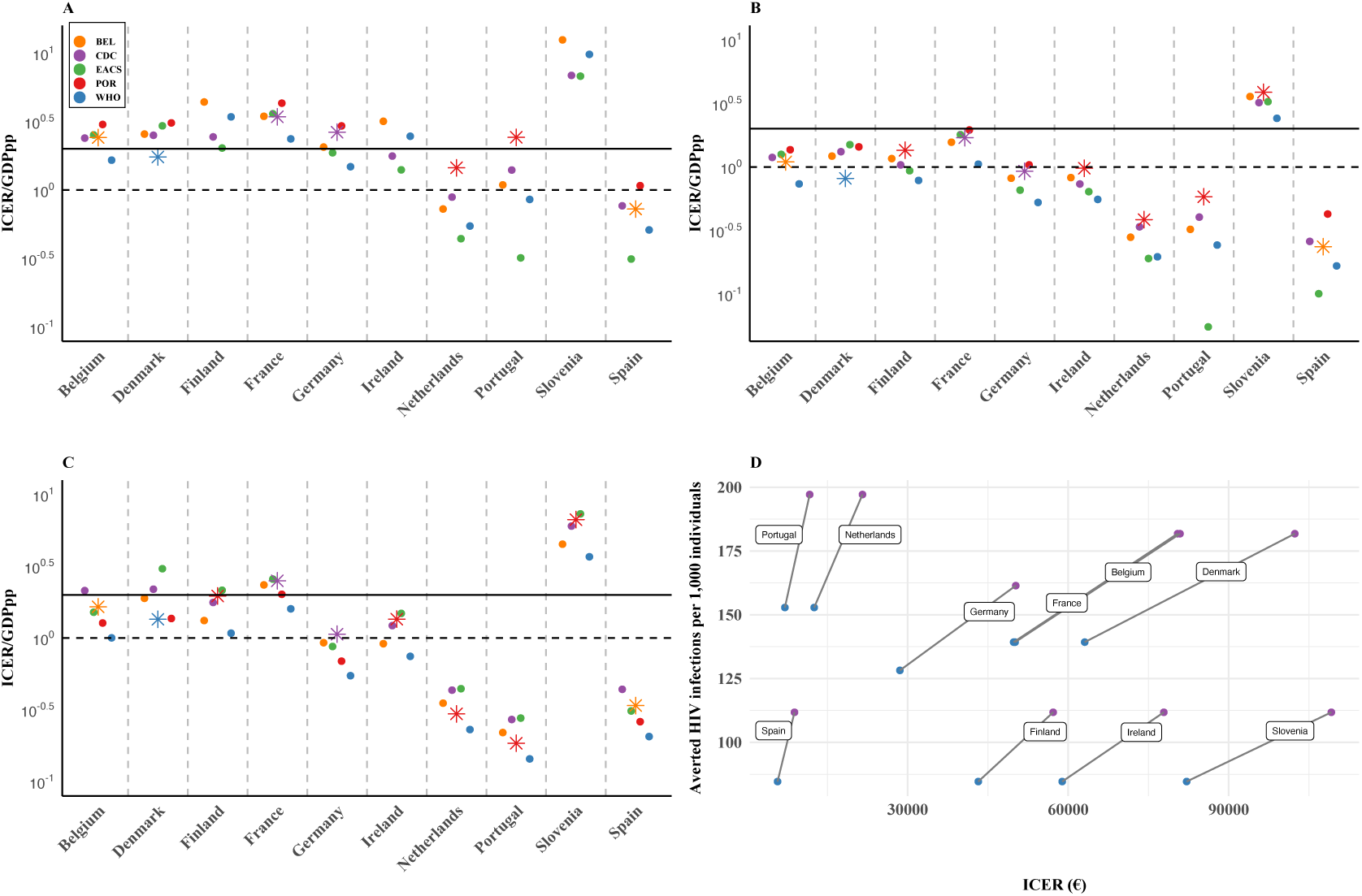
Cost-effectiveness of the different PrEP policies in the 10 countries currently reimbursing PrEP. (A) ICER over GDP per person at the 10-year time horizon; (B) ICER over GDP per person at the 20-year time horizon; (C) ICER over GDP per person at the 20-year time horizon, assuming that each country keeps the current policy for 5 years and then switches to the selected policy. In (A), (B) and (C) the solid horizontal line corresponds to an ICER of twice the GDP per person. (D) Comparison between the most effective policy (CDC) and the most cost-effective policy (WHO) in terms of ICER and averted infection rates. The slope of the gray lines indicates measure the change in averted infections per unit increase in ICER.

**Table 3:**
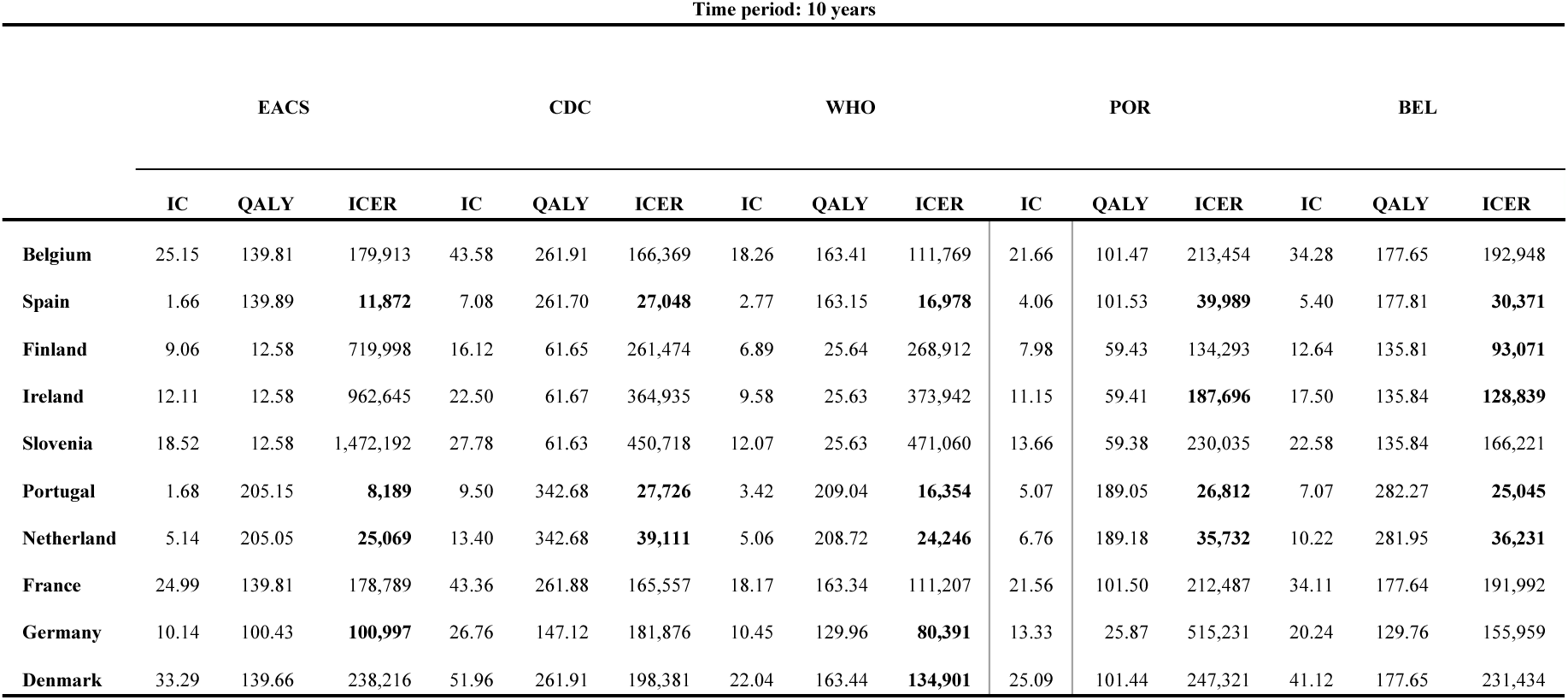
Cost-effectiveness evaluation of the different PrEP policies in the countries reimbursing PrEP, at the 10-year time horizon. IC is incremental cost, expressed in M€ (million Euros). QALY is the number of QALY gained. ICER is the Incremental Cost-Effectiveness Ratio, expressed in €/QALY (Euros per QUALY gained). Bold ICER means that cost-effectiveness is achieved at the threshold of twice the GDP per person.

**Table 4:**
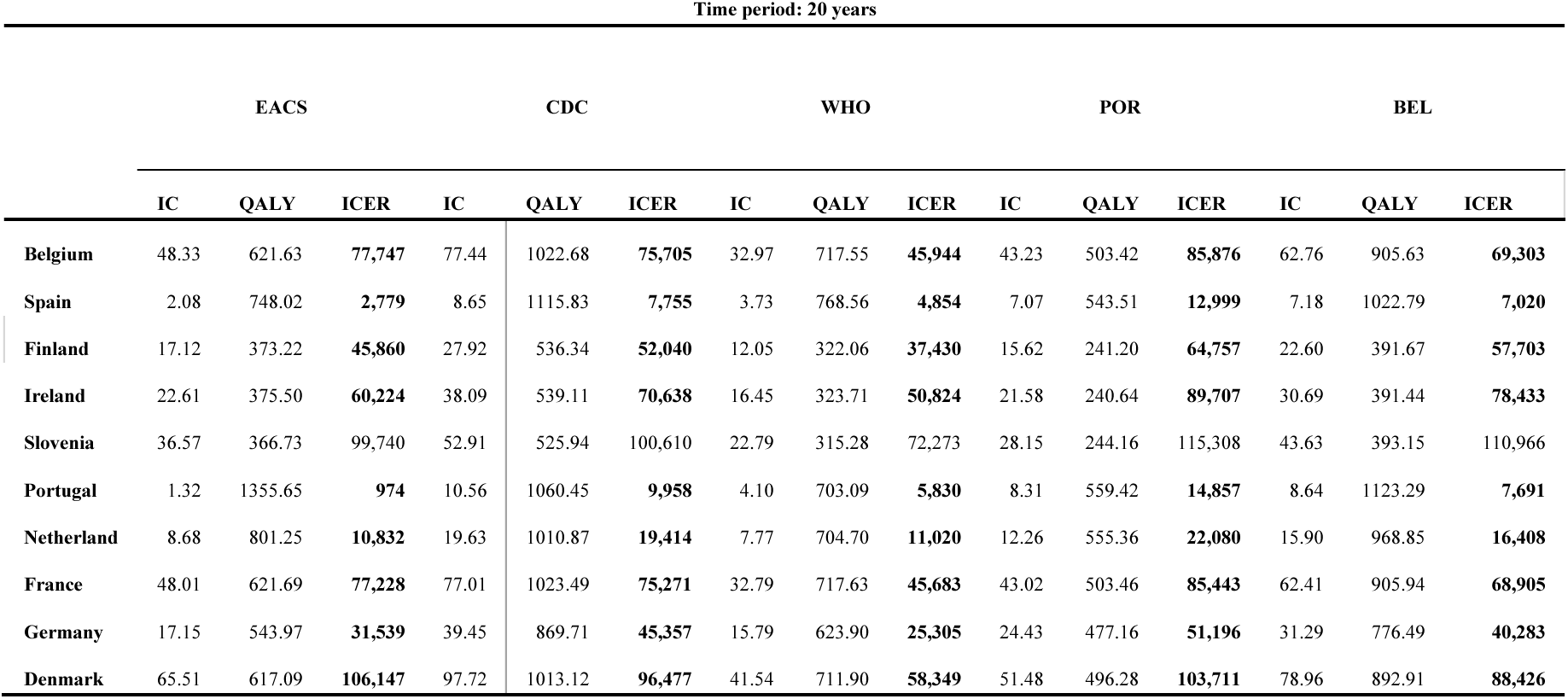
Cost-effectiveness evaluation of the different PrEP policies in the countries reimbursing PrEP, at the 20-year time horizon. IC is incremental cost, expressed in M€ (million Euros). QALY is the number of QALY gained. ICER is the Incremental Cost-Effectiveness Ratio, expressed in €/QALY (Euros per QUALY gained). Bold ICER means that cost-effectiveness is achieved at the threshold of twice the GDP per person.

Figure 3B reports ICER at the time horizon of 20 years, and we observed that all the examined policies were cost-effective in all the countries which currently reimburse PrEP, except in Slovenia, where still none of the policies were cost-effective. Again, the currently implemented policy was the most cost-effective only in Denmark (WHO), and the most cost-effective policies were either WHO or EACS. WHO was the most cost-effective policy in all the countries where it was already so at the 10-year horizon. EACS was still the most cost-effective in the Netherlands (albeit tied with WHO), Portugal and Spain, but it was no longer so in Finland and Slovenia, replaced by WHO. However, it is worth noting that in both countries WHO had been among the least cost-effective at the 10-year horizon.

In Figures 3A and 3B, we assumed that each country immediately switched to each examined policy, if not already adopted. Instead, in Figure 3C we examine the scenario in which a country kept the current policy for five years and then transitioned to another policy. Describing the results, we refer to the policy adopted after the five-year period. No policy was cost-effective in Slovenia, and all policies were cost-effective everywhere except for EACS in Denmark, Finland and France, for CDC in Belgium, Denmark and France, and for BEL in France. WHO was the most cost-effective policy everywhere, tied with POR in Denmark.

In Figure 3D we compare the CDC policy - the most effective policy in averting infections - with the WHO policy – the most cost-effective policy emerging from Figure 3C -- at the 20-year horizon. We show ICER vs averted infection and the slope of the line connecting the two policies indicates the additional number of HIV infections averted per unit increase in the ICER. The Netherlands, Portugal and Spain feature steep slopes, meaning that the rate of averted infections beyond the most cost-effectiveness policy would entail a limited increase in costs. In opposition, flatter slopes like in Ireland and Slovenia signal that increasing the rate of averted infections beyond the most cost-effectiveness policy would entail significantly higher costs.

Finally, in Appendix pp 8 we show the effect of different economic assumptions through a sensitivity analysis. Yearly discounting rate and ART cost appear as the two factors most impacting cost-effectiveness estimates.

### Health economics analysis in countries not reimbursing PrEP

We estimated the willingness-to-pay (WTP) threshold for each policy, in the countries which currently do not reimburse PrEP, at the 20-year horizon. The WTP threshold is the maximum cost of PrEP for which a policy is cost-effective. Any cost at or below the WTP threshold causes the policy to be cost-effective. We estimated the WTP threshold in the countries under study and compared it to the reported cost of generic PrEP, and of Truvada in the case of Hungary and Italy, where it is available.In Figure 4 we present our estimations and we can observe that the cost of Truvada exceeded the WTP threshold of any policy by more than 100 Euros per week, making it extremely not cost-efficient. On the contraty, all the tested policies were cost-effective at the current cost of generic PrEP in Cyprus and Italy, while none was cost-effective in Bulgaria and Hungary. BEL and WHO policies were cost-effective in all the remaining countries, while POR was cost-effective in Lithuania and Romania, and CDC and EACS in Greece and Latvia.

**Figure 4.**
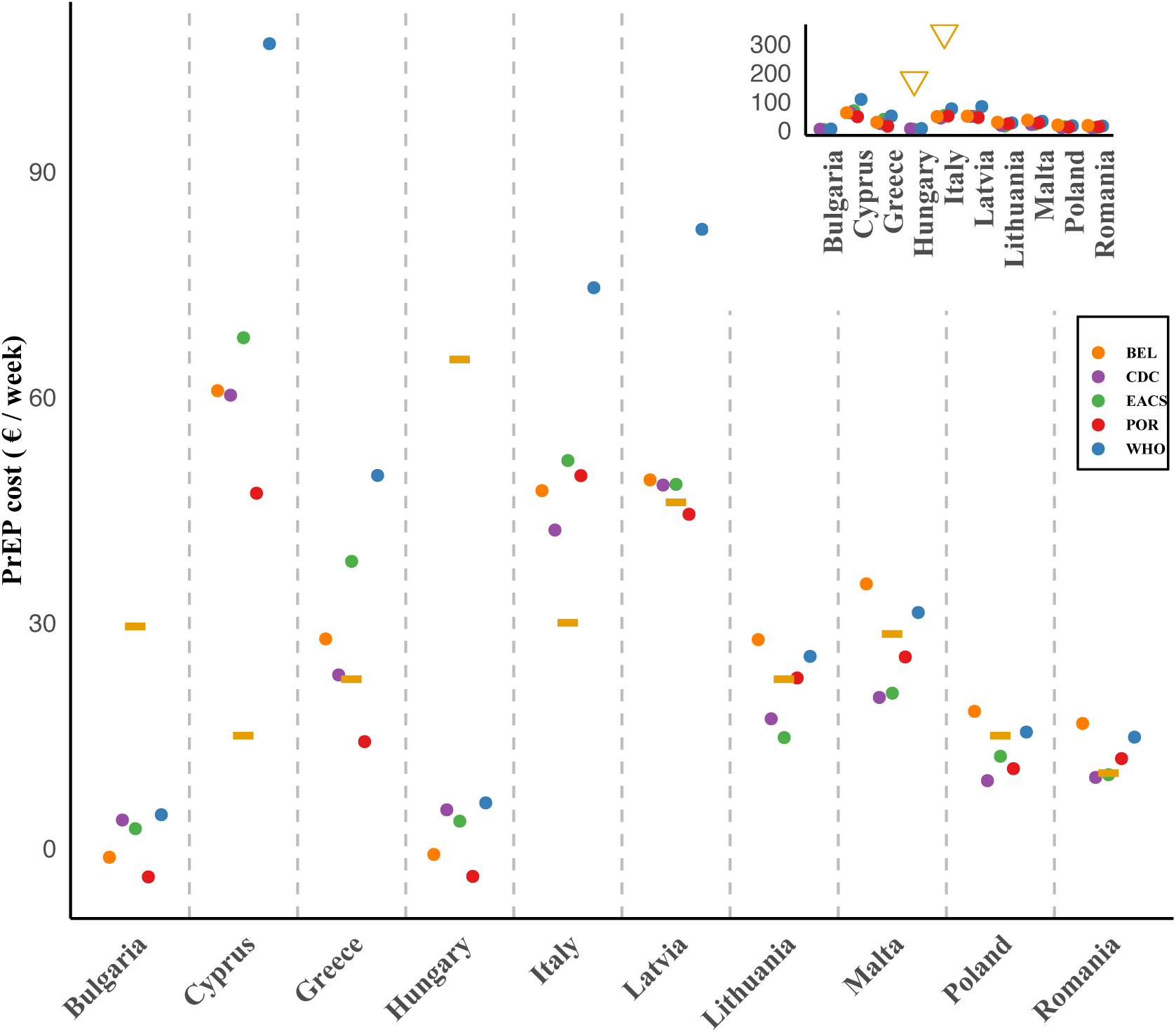
Willingness-to-pay threshold of each examined PrEP policy in the countries not reimbursing PrEP, vs the currently estimated cost of generic PrEP (yellow bar) and Truvada (yellow triangle). The main plot excludes Truvada, the inset includes Truvada.

## 5 Discussion

Our study evaluated the effectiveness and cost-effectiveness of alternative policies for the supply of PrEP for HIV to men who have sex with men (MSM) in 20 countries of the EU, over the next 20 years. We used a stochastic agent-based model for HIV infection and we estimated the effectiveness of PrEP policies in terms of averted infections and HIV-related deaths. In our estimations, all examined PrEP policies substantially decreased infections and deaths, except in communities where HIV prevalence was very low (below 5%). The guidelines proposed by the CDC and BEL policies exhibited the highest effectiveness, across all examined countries. Their performance is clearly attributable to the fact that they entailed a very high PrEP coverage. Nonetheless, the WHO guidelines achieved a moderately lower effectiveness than CDC and BEL, but at a substantially lower PrEP coverage. This clearly required a comparison among policies that accounted for their economic viability as well as effectiveness.

Over a 20-year horizon, we observed that, barring extreme conditions (exceptional high PrEP costs coupled with low ART costs), PrEP interventions under any policy were cost-effective when compared to the benchmark of twice the GDP per capita. This proves that in the EU, integrating PrEP in HIV prevention strategies among MSM should be economically sustainable across countries and public health approaches. This is consistent with previous findings in the Netherlands as well as other high-income regions like the US, UK, and Australia(8,37,40,41). Moreover, we found that no country, except Denmark, is now adopting the most cost-effective strategy. This suggests the lack of a quantitative assessment of the effectiveness and cost-effectiveness of current strategies (if any) and of alternative ones, to inform possible policy adjustments that are compatible with national and global prevention targets, as well as economic concerns. We also noted that the WHO policy was consistently the most cost-effective, both in the shorter and longer term. This is compatible with the fact that targeted PrEP strategies may be preferable in high-income settings(23). In practice, our analyses show that the WHO policy is likely to be suitable to those countries, like Slovenia, where the cost of PrEP is high, and thus deviating from optimal cost-effectiveness is expensive. This is true both as an initial choice and a policy to switch to: Countries which currently do not provide PrEP free-of-charge could start providing it following the WHO guidelines, and countries which already provide PrEP and are following other policies might benefit from transitioning to WHO’s policy.

In countries where instead PrEP is cheaper, such as Spain, Portugal and the Netherlands, we showed that policies that achieve high PrEP coverage (CDC and BEL) are appropriate, as they maximize the number of averted infections and deaths at a limited additional cost (. We remark that these policies are also appropriate where maximizing the number of averted infections may outweigh economic considerations, as it is likely to be currently the case in France and Germany. In this context, our results also suggest that those countries should not switch to WHO unless cost saving becomes pressing, as this would reduce effectiveness.

Additionally, as expected, we found that the national cost of PrEP is an important factor in determining the cost-effectiveness of each policy in countries which do not yet provide PrEP free of charge. Specifically, the cost of Truvada showed to be substantially higher than the willingness-to-pay threshold of any policy, signaling that it is unlikely to be part of any strategy if economic concerns are present. The cost of generic PrEP, instead, could be above or below the threshold depending on the country and the policy adopted. This shows that lowering procurement costs is a crucial part of PrEP policy design, as it would decrease the marginal cost of increasing PrEP coverage, giving countries the option to select strategies that prioritize effectiveness. Our sensitivity analysis also showed that cost-effectiveness is sensitive to the chosen discounting rate. We thus recommend future updates of the cost-effectiveness estimates in the case of major shifts in inflationary trends.

Our study has limitations. First, cost estimates focused on PrEP and ART, and may underestimate the total cost of PrEP policies. Notwithstanding, our conservative estimates consistently demonstrated the cost-effectiveness of PrEP interventions. Second, to inform HIV prevalence among MSM, we used reported rates of HIV diagnosis, which may be biased where testing rates are low or unknown (33). Third, we deliberately excluded sexually transmitted infections from the analysis. The purpose of PrEP is to protect from HIV and we believe that it is useful to assess its effectiveness and cost-effectiveness on the basis of its impact on HIV infections and related negative health outcomes. In addition, both the evidence of the effect of PrEP on the spread of other STI is mixed(8,11) and the circulation of STI in Europe among MSM is poorly known(42). Including STI in the analysis would thus inevitably increase the uncertainty of our predictions, without informing on what factors really determine the effectiveness and cost-effectiveness of policies. Finally, our work is restricted to oral PrEP. Future studies should update the cost-effectiveness of the examined policies to include alternative formulations as they become available in the EU, and their costs becomes clear.

## Data Availability

All data produced are available online at https://github.com/ev-modelers

## References

1. Palich R, Arias-Rodríguez A, Duracinsky M, Le Talec JY, Rousset TO, Lascoux-Combe C, et al. High proportion of post-migration HIV acquisition in migrant men who have sex with men receiving HIV care in the Paris region, and associations with social disadvantage and sexual behaviours: results of the ANRS-MIE GANYMEDE study, France, 2021 to 2022. Euro Surveill. 2024;29(11):2300445.

2. Joint United Nations Programme on HIV/AIDS (UNAIDS). The Urgency of Now: AIDS at a Crossroads [Internet]. 2024. Available from: https://www.unaids.org/en/resources/documents/2024/global-aids-update-2024

3. Grant RM, Lama JR, Anderson PL, McMahan V, Liu AY, Vargas L, et al. Preexposure Chemoprophylaxis for HIV Prevention in Men Who Have Sex with Men. N Engl J Med. 2010 Dec 30;363(27):2587–99.

4. Molina JM, Capitant C, Spire B, Pialoux G, Cotte L, Charreau I, et al. On-Demand Preexposure Prophylaxis in Men at High Risk for HIV-1 Infection. N Engl J Med. 2015 Dec 3;373(23):2237– 46.

5. McCormack S, Dunn DT, Desai M, Dolling DI, Gafos M, Gilson R, et al. Pre-exposure prophylaxis to prevent the acquisition of HIV-1 infection (PROUD): effectiveness results from the pilot phase of a pragmatic open-label randomised trial. The Lancet. 2016 Jan 2;387(10013):53–60.

6. Jourdain H, others. Real-world effectiveness of pre-exposure prophylaxis in men at high risk of HIV infection in France: a nested case-control study. Lancet Public Health. 2022;7(6):e529–36.

7. Molina JM, Ghosn J, Assoumou L, Delaugerre C, Algarte-Genin M, Pialoux G, et al. Daily and on-demand HIV pre-exposure prophylaxis with emtricitabine and tenofovir disoproxil (ANRS PREVENIR): a prospective observational cohort study. Lancet HIV. 2022 Aug 1;9(8):e554–62.

8. Traeger MW, Guy R, Asselin J, Patel P, Carter A, Wright EJ, et al. Real-world trends in incidence of bacterial sexually transmissible infections among gay and bisexual men using HIV pre-exposure prophylaxis (PrEP) in Australia following nationwide PrEP implementation: an analysis of sentinel surveillance data. Lancet Infect Dis. 2022 Aug;22(8):1231–41.

9. Gökengin D, Noori T, Alemany A, Bienkowski C, Liegon G, İnkaya AÇ, et al. Prevention strategies for sexually transmitted infections, HIV, and viral hepatitis in Europe. Lancet Reg Health – Eur [Internet]. 2023 Nov 1 [cited 2024 Sep 11];34. Available from: https://www.thelancet.com/journals/lanepe/article/PIIS2666-7762(23)00157-6/fulltext

10. Grov C, Westmoreland DA, D’Angelo AB, Pantalone DW. How Has HIV Pre-Exposure Prophylaxis (PrEP) Changed Sex? A Review of Research in a New Era of Bio-behavioral HIV Prevention. J Sex Res. 2021 Sep 2;58(7):891–913.

11. Protiere C, Sagaon-Teyssier L, Donadille C, Sow A, Gaubert G, Girard G, et al. Perception of PrEP-related stigma in PrEP users: Results from the ANRS-PREVENIR cohort. HIV Med. 2023;24(8):938–45.

12. Castro DR, Delabre RM, Molina JM. Give PrEP a chance: moving on from the “risk compensation” concept. J Int AIDS Soc. 2019;22(S6):e25351.

13. Health Programme of the European Union, AIDS Action Europe. Rapid Assessment on Access to PrEP in EU/EEA Countries, 2022. 2023 Mar.

14. Meireles P, Plankey M, Rocha M, Rojas J, Brito J, Barros H. Eligibility for Pre-exposure Prophylaxis According to Different Guidelines in a Cohort of HIV-Negative Men Who Have Sex with Men in Lisbon, Portugal. Sex Res Soc Policy. 2020 Dec;17(4):688–99.

15. Meireles P, Plankey M, Rocha M, Brito J, Mendão L, Barros H. Different guidelines for pre-exposure prophylaxis (PrEP) eligibility estimate HIV risk differently: an incidence study in a cohort of HIV-negative men who have sex with men, Portugal, 2014-2018. Euro Surveill Bull Eur Sur Mal Transm Eur Commun Dis Bull. 2020 Jul;25(28):1900636.

16. Brunner P, Brunner K, Kübler D. The Cost-Effectiveness of HIV/STI Prevention in High-Income Countries with Concentrated Epidemic Settings: A Scoping Review. AIDS Behav. 2022 Jul 1;26(7):2279–98.

17. Nichols BE, Boucher CAB, Valk M van der, Rijnders BJA, Vijver DAMC van de. Cost-effectiveness analysis of pre-exposure prophylaxis for HIV-1 prevention in the Netherlands: a mathematical modelling study. Lancet Infect Dis. 2016 Dec 1;16(12):1423–9.

18. van de Vijver DAMC, Richter AK, Boucher CAB, Gunsenheimer-Bartmeyer B, Kollan C, Nichols BE, et al. Cost-effectiveness and budget effect of pre-exposure prophylaxis for HIV-1 prevention in Germany from 2018 to 2058. Eurosurveillance. 2019 Feb 14;24(7):1800398.

19. Nurchis MC, Riccardi MT, Sapienza M, Pascucci D, Damiani G. Moving forward: Cost-Effectiveness of PrEP in HIV prevention for Men Who Have Sex with Men in Italy. Eur J Public Health. 2020 Sep 1;30(Supplement_5):ckaa165.994.

20. Ousseine YM, Lydié N, Velter A. Pre-exposure prophylaxis in France: How many MSM are eligible and how much will it cost? PLOS ONE. 2022 dic;17(12):e0278016.

21. Bozzani FM, Terris-Prestholt F, Quaife M, Gafos M, Indravudh PP, Giddings R, et al. Costs and Cost-Effectiveness of Biomedical, Non-Surgical HIV Prevention Interventions: A Systematic Literature Review. PharmacoEconomics. 2023 May 1;41(5):467–80.

22. Babel RA, Wang P, Alessi EJ, Raymond HF, Wei C. Stigma, HIV Risk, and Access to HIV Prevention and Treatment Services Among Men Who have Sex with Men (MSM) in the United States: A Scoping Review. AIDS Behav. 2021 Nov;25(11):3574–604.

23. Steinegger B, Iacopini I, Teixeira AS, Bracci A, Casanova-Ferrer P, Antonioni A, et al. Non-selective distribution of infectious disease prevention may outperform risk-based targeting. Nat Commun. 2022 May 31;13(1):3028.

24. Jenness SM, Goodreau SM, Morris M. EpiModel: An R Package for Mathematical Modeling of Infectious Disease over Networks. J Stat Softw [Internet]. 2018 Apr [cited 2024 Sep 12];84. Available from: https://www.ncbi.nlm.nih.gov/pmc/articles/PMC5931789/

25. Neumann PJ, Sanders GD, Russell LB, Siegel JE, Ganiats TG, editors. Cost-Effectiveness in Health and Medicine [Internet]. New York: Oxford University Press; 2016. Available from: https://academic.oup.com/book/12265

26. World Health Organization WH. WHO implementation tool for pre-exposure prophylaxis (PrEP) of HIV infection: module 1: clinical [Internet]. World Health Organization; 2017 [cited 2024 Sep 11]. Report No.: WHO/HIV/2017.17. Available from: https://iris.who.int/handle/10665/255889

27. Georg Behrens, AP. European AIDS Clinical Society Guidelines Guidelines 9.0 [Internet]. European AIDS Clinical Society (EACS); 2017 Oct. Available from: https://www.eacsociety.org/guidelines/eacs-guidelines/

28. CDC. Centers for Disease Control and Prevention: US Public Health Service: Preexposure prophylaxis for the prevention of HIV infection in the United States—2017 Update: a clinical practice guideline. [Internet]. 2018 Mar. Available from: https://www.cdc.gov/hiv/pdf/risk/prep/cdc-hiv-prep-guidelines-2017.pdf

29. Baetselier ID, Reyniers T, Nöstlinger C, Wouters K, Fransen K, Crucitti T, et al. Pre-Exposure Prophylaxis (PrEP) as an Additional Tool for HIV Prevention Among Men Who Have Sex With Men in Belgium: The Be-PrEP-ared Study Protocol. JMIR Res Protoc. 2017 Jan 30;6(1):e6767.

30. Jenness SM, Maloney KM, Smith DK, Hoover KW, Goodreau SM, Rosenberg ES, et al. Addressing Gaps in HIV Preexposure Prophylaxis Care to Reduce Racial Disparities in HIV Incidence in the United States. Am J Epidemiol. 2019 Apr 1;188(4):743–52.

31. Hamilton DT, Agutu C, Sirengo M, Chege W, Goodreau SM, Elder A, et al. Modeling the impact of different PrEP targeting strategies combined with a clinic-based HIV-1 nucleic acid testing intervention in Kenya. Epidemics. 2023 Sep 1;44:100696.

32. Robineau O, Velter A, Barin F, Boelle PY. HIV transmission and pre-exposure prophylaxis in a high risk MSM population: A simulation study of location-based selection of sexual partners. PLOS ONE. 2017 Nov 30;12(11):e0189002.

33. Marcus U, Hickson F, Weatherburn P, Schmidt AJ, Network E. Prevalence of HIV among MSM in Europe: comparison of self-reported diagnoses from a large scale internet survey and existing national estimates. BMC Public Health. 2016;16(1):1–10.

34. Rocha M, Meireles P, Esteves F, Areal A, Barros H, Martins MR. Sexual behavior and partner network parameters from the Lisbon cohort study. Sci Data. 2020;7(1):1–9.

35. Morris M, Kretzschmar M. Concurrent partnerships and the spread of HIV. AIDS. 1997;11(5):641–8.

36. ECDC. Pre-exposure prophylaxis for HIV prevention in Europe and Central Asia [Internet]. Stockholm; 2023. Available from: https://www.ecdc.europa.eu/en/publications-data/hiv-infection-prevention-pre-exposure-prophylaxis-monitoring-dublin

37. Cambiano V, Miners A, Dunn D, McCormack S, Ong KJ, Gill ON, et al. Cost-effectiveness of pre-exposure prophylaxis for HIV prevention in men who have sex with men in the UK: a modelling study and health economic evaluation. Lancet Infect Dis. 2018 Jan;18(1):85–94.

38. Bank W, OECD. World Bank national accounts data, and OECD National Accounts data files. 2019.

39. Husereau D, Drummond M, Augustovski F, De Bekker-Grob E, Briggs AH, Carswell C, et al. Consolidated Health Economic Evaluation Reporting Standards (CHEERS) 2022 Explanation and Elaboration: A Report of the ISPOR CHEERS II Good Practices Task Force. Value Health. 2022 Jan;25(1):10–31.

40. Boerekamps A, van Sighem A, van Agtmael M, Claessen F, Richter C, Sprangers M, et al. Cost-effectiveness of pre-exposure prophylaxis (PrEP) in preventing HIV-1 infections in the Netherlands: a mathematical modelling study. PLoS One. 2018;13(3):e0194200.

41. Wheatley MM, Knowlton G, Kao SY, Jenness SM, Enns EA. Cost-Effectiveness of Interventions to Improve HIV Pre-exposure Prophylaxis Initiation, Adherence, and Persistence Among Men Who Have Sex With Men. J Acquir Immune Defic Syndr. 2022;90(1).

42. European Centre for Disease Prevention and Control. Rising rates of sexually transmitted infections across Europe [Internet]. 2023. Available from: https://www.ecdc.europa.eu/en/news-events/rising-rates-sexually-transmitted-infections-across-europe

